# Mitochondrial and Stress-Related Psychobiological Regulation of FGF21 in Humans

**DOI:** 10.1101/2025.01.30.25321437

**Authors:** Mangesh Kurade, Natalia Bobba-Alves, Catherine Kelly, Alexander Behnke, Quinn Conklin, Robert-Paul Juster, Michio Hirano, Caroline Trumpff, Martin Picard

## Abstract

FGF21 is a metabolic hormone induced by fasting, metabolic stress, and mitochondrial oxidative phosphorylation (OxPhos) defects that cause mitochondrial diseases (MitoD). Here we report that acute psychosocial stress alone (without physical exertion) decreases serum FGF21 by an average of 20% (*p*<0.0001) in healthy controls but increases FGF21 by 32% (*p*<0.0001) in people with MitoD—pointing to a functional interaction between the stress response and OxPhos capacity in regulating FGF21. We further define co-activation patterns between FGF21 and stress-related neuroendocrine hormones and report novel associations between FGF21 and psychosocial factors related to stress and wellbeing, highlighting a potential role for FGF21 in meeting the energetic needs of acute and chronic psychosocial stress.

## Main Text

The hormone fibroblast growth factor 21 (FGF21) regulates systemic energy metabolism [1]. It is released by the liver and other organs in response to metabolic challenges, including fasting, and acts on the brain and multiple tissues in the periphery to regulate nutrient intake and metabolic processes [2-5]. **Figure S1** and **Table S1** show RNA transcript levels for FGF21 (highest in liver, muscle, and immune cells) and its receptor FGFR1 (highest in arteries, pancreas, and small intestine) across 50 human tissues—establishing their expression across several organs systems subject to psychobiological processes.

FGF21 is considered a relatively specific and sensitive biomarker of pediatric and adult mitochondrial diseases (MitoD) caused by genetic oxidative phosphorylation (OxPhos) defects: it is elevated in most clinical cases [6, 7] and in several animal models of MitoD [3, 4], illustrating the relevance of mitochondrial OxPhos capacity and energy metabolism in regulating FGF21 biology. In animals, mitochondrial OxPhos capacity and energy metabolism are also linked to mental and psychosocial states [8] because perceived threats trigger endocrine responses (e.g., cortisol, norepinephrine) that induce potent physiological recalibrations (e.g., increased heart rate and blood pressure) that mobilize metabolic resources to fuel energetically demanding “fight-or-flight” stress responses [9, 10]. Interestingly, some animal models of chronic stress [11] and human studies of psychiatric illnesses [12, 13] have reported elevated FGF21 levels. These findings lead us to reason that mitochondrial OxPhos defects and psychosocial factors may therefore converge to regulate FGF21 biology in humans.

To probe this question, we enrolled three groups of participants as part of the Mitochondrial Stress, Brain Imaging, and Epigenetics (MiSBIE) study [14]: individuals with normal mitochondrial biology (*Controls, n*=70); individuals with a mitochondrial DNA mutation (m.3243A>G, *Mutation, n*=25, including 5 individuals with the most severe form of MitoD known as mitochondrial encephalomyopathy, lactic acidosis, stroke-like episodes [*MELAS*]); and individuals with a single, large-scale mtDNA deletion (*Deletion, n*=15) causing MitoD. Detailed information about the study procedures and measures can be found the *Supplemental Methods* and in [14].

We first confirmed two hallmarks of FGF21 biology: its induction by OxPhos defects and its link to fasting and systemic metabolism. Participants arrived at 09:00AM and completed anthropomorphic measurements and a fasting blood draw at ∼10:00AM; after which they received a standardized breakfast, completed questionnaires, and remained relatively sedentary before consuming a standardized lunch. Blood was collected after lunch at ∼1:00PM (13:00) to assess FGF21 levels in the fed state. Compared to controls, MitoD participants exhibited markedly elevated serum FGF21 levels (180-420% higher, *ps*<0.0001, **Figure 1a)**, which were highest in the Deletion group (**Figure S2a**). In line with the inducibility of FGF21 by fasting, AM-fasted FGF21 concentrations were higher than PM-fed concentrations across all groups (*p*s<0.001, **Figure S2a-c**). This morning-to-afternoon decline was more pronounced in Controls (Δ-73.3%, *g=*0.5, *p<*0.0001) than in MitoD (Δ-14.7%, *g=*0.1, *p<*0.0005; **Figure S2c**), providing additional evidence that FGF21 regulation is altered in MitoD. Using a series of ROC curve analyses, we also confirmed that FGF21 is a biomarker of human OxPhos deficiency with sensitivity and specificity characteristics comparable with prior studies—noting that FGF21 measured in the fed state was most predictive of MitoD (**Figure S3a-d**).

**Figure 1.**
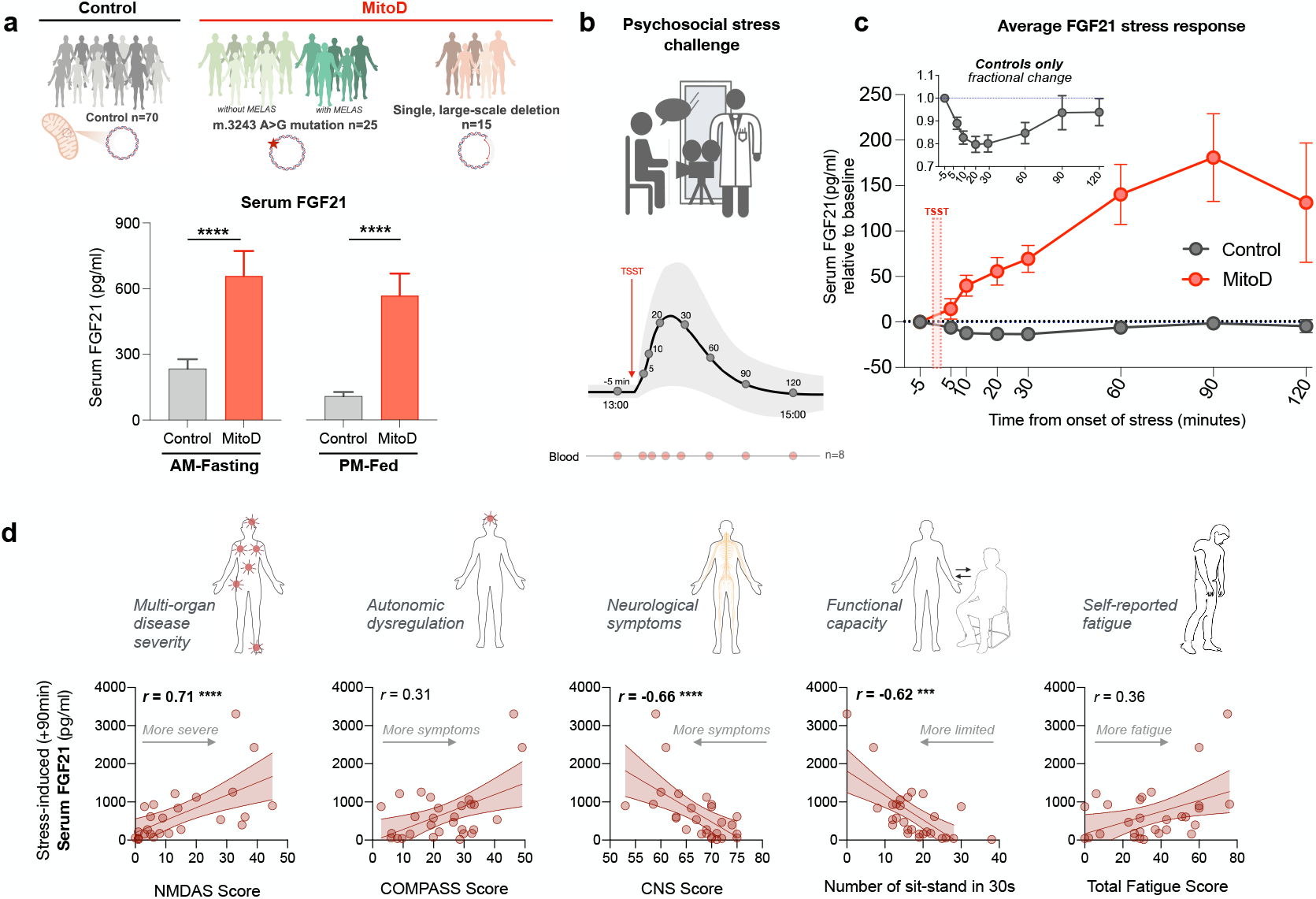
Elevated serum FGF21 in mitochondrial disease shows abnormal stress response and correlates with disease severity. **(a)** Distribution of the study cohorts and Mann-Whitney’s *U*-tests comparing mean serum FGF21 levels in controls and MitoD when fasted (AM) (Controls: 235.0 pg/ml, *n*=65 vs MitoD: 657.8 pg/ml, *n*=34) or fed (PM) (Controls: 109.0 pg/ml, *n*=65 vs MitoD: 567.8 pg/ml, *n*=33). Error bars represent SEM. **(b)** Schematic of the modified TSST paradigm used to assess FGF21 and hormone responses to acute stress. **(c)** Changes in serum FGF21 levels (pg/ml) in response to the socio-evaluative stressor in controls (*n*=60–65) and MitoD (*n*=33–28). Values at each time point are expressed as absolute change (pg/ml) relative to baseline measurements at -5 min (Absolute change = Value – Baseline). *Inset:* Relative change in serum FGF21 (pg/ml) within controls relative to baseline measurement (Relative change = Value/Baseline). **(d)** Spearman correlations between disease severity scores and serum FGF21 expression at 90-min post-stress in the MitoD cohort. Two-tailed significance is indicated as *p<*0.05 (*), *p<*0.01 (**), *p<*0.001 (***), *p<*0.0001 (****).

Consistent with some known factors linked to FGF21 regulation [15, 16], fasting FGF21 in controls was comparable between females and males and was positively correlated with age (*r*_*s*_*=*0.37, *p=*0.002), percent body fat (*r*_s_*=*0.25, *p=*0.046), total fat mass (*r*_s_*=*0.31, *p=*0.015), fasting blood glucose (*r*_s_*=*0.49, *p<*0.0001), insulin (*r*_s_=0.027, *p=*0.026), and total cholesterol (*r*_s_*=*0.30, *p=*0.016), but negatively correlated with HDL (*r*_s_*=*-0.35, *p=*0.0043) (**Figures S4a-d**). Apart from the correlation between FGF21 and percent body fat (*r*_s_*=*0.42, *p=*0.013), these relationships were not observed in the MitoD group, indicating that OxPhos defects disproportionally elevate FGF21 such that these relationships are obscured compared to healthy controls.

To isolate the influence of psychological stress without the confounding effects of exertion or hunger [17], participants performed an experimental socio-evaluative stress test beginning at ∼13:00 in the afternoon. The stressor consisted of 2 minutes of mental preparation and a 3-minute speech delivery in front of an “evaluator” wearing a white coat—a widely used laboratory stressor that reliably elicits psychophysiological reactivity (see *Methods* and [14] for details). Serum FGF21 was measured in blood drawn through an intravenous catheter immediately before the onset of the stressor (-5) and at +5, 10, 20, 30, 60, 90, and 120 minutes after the stressor, for a total of 8 afternoon timepoints, allowing us to establish for the first time the acute reactivity of FGF21 to psychosocial stress both in healthy controls and MitoD patients (**Figure 1b**).

A mixed effects model demonstrated that the stress-induced FGF21 response was moderated by mitochondrial OxPhos defects, with Control and MitoD participants exhibiting distinct FGF21 trajectories (**Figure S5**). FGF21 levels in controls decreased by an average of - 20% (Δ -13.10 pg/ml, *g=*0.75, *p*<0.0001) 20 minutes after the onset of the stressor, and had essentially recovered by the 90-min timepoint (**Figure 1c**, *inset*, and **Figure S6a**). By contrast, FGF21 levels in MitoD patients *increased*, peaking at 90 minutes with an average change of +32% (Δ+180.8 pg/ml, *g=*1.17, *p<*0.0001, **Figure 1c**). All MitoD subgroups (Mutation with or without MELAS, and Deletion) showed similar trajectories (**Figure S6b**).

To explore the importance of FGF21 as a clinical marker, we evaluated how serum FGF21 levels—sampled during fasting, after lunch, and under stress—related to important functional and clinical markers of mitochondrial disease, including i) clinician-assessed disease severity, ii) objective tests of functional capacity, and iii) patient-reported fatigue. MitoD individuals with higher peak FGF21 (measured 90 minutes post-stress) had more severe disease manifestations overall (|*r*_s_|*=*0.66–0.71, **Figure 1d**). Interestingly, these FGF21-disease associations were always *stronger* than those observed with the AM-fasted values (**Figure S7a and b**). These findings suggest that mental stress evokes robust and clinically meaningful changes (rather than noise) in circulating FGF21 in humans, pointing to shared psychobiological processes linking acute mental stress and OxPhos defects.

To investigate the potential basis for the differential effects of acute psychosocial stress on FGF21 dynamics between controls and MitoD, we examined the interactions between FGF21 and other hormones of the hypothalamic-pituitary-adrenal (HPA), hypothalamic-pituitary-gonadal (HPG), and sympathetic nervous system (SNS) axes, which were measured from blood and saliva across the same afternoon time course.

Compared to controls, MitoD patients demonstrated distinct correlation patterns between FGF21 and stress-induced steroid hormones involved in the HPA (i.e., cortisol and cortisone) and HPG (i.e., DHEA and testosterone) axes across the stress reactivity-recovery time course (**Figure 2a**). For cortisol and cortisone, controls exhibited no correlation or small negative correlations with FGF21 across timepoints, whereas the MitoD group displayed moderate to large positive correlations across most timepoints. Correlations between DHEA and FGF21 followed a similar pattern, with mostly negative correlations across timepoints in controls, alongside mostly positive correlations in MitoD. This pattern was reversed for testosterone and FGF21: controls showed a mix of small, positive and negative correlations across time, while the MitoD group showed significant negative correlations across all timepoints. These data reveal differential co-regulation patterns between FGF21 and anabolic/catabolic endocrine axes between individuals with normal or abnormal mitochondrial OxPhos.

**Figure 2.**
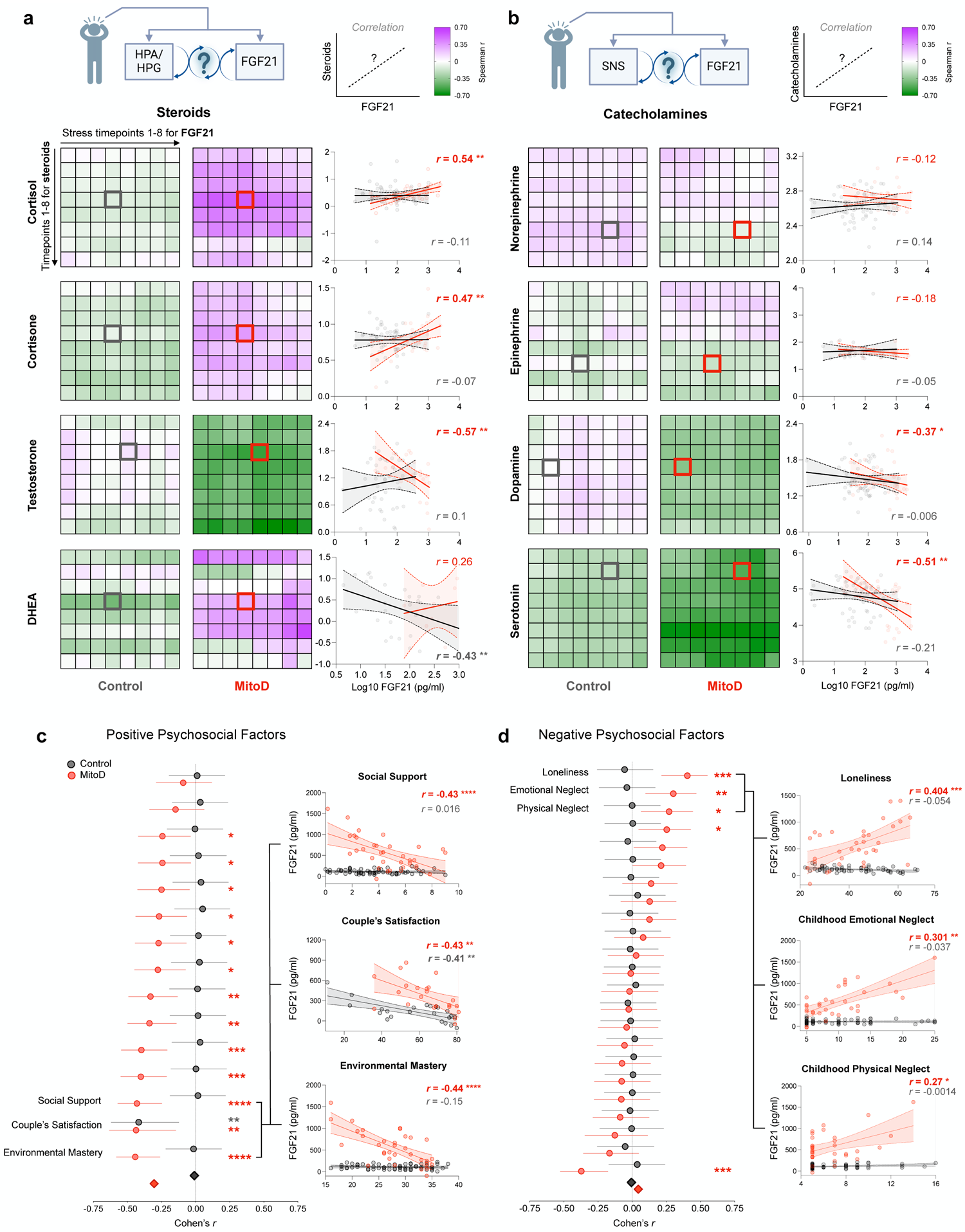
Patterns of co-regulation between FGF21 and stress hormones, and FGF21 and psychosocial factors differ in controls and patients with Mitochondrial Disease. *Top:* Spearman correlation matrices depicting differential patterns in controls and MitoD patients’ stress-induced co-regulation patterns between FGF21 and corresponding measurements of **(a)** steroid hormones of the HPA and HPG axes (Cortisol: *n*=27–62, Cortisone: *n*=27–63, Testosterone: *n*=20–52, DHEA: *n*=20–58), and **(b)** hormones involved in the SNS (Epinephrine: *n*=15–38, Norepinephrine: *n*=29–64, Dopamine: *n*=29–64) measured across the TSST time course. All measurements are Log_10_ transformed. Scatter plots depict correlations at specific time points featuring moderate to high correlation in MitoD compared to their counterparts in controls. Linear estimates are provided for illustrative purposes. *Bottom:* Forrest plots showing associations between FGF21 levels, adjusted for age and percent fat, and psychosocial self-report measures categorized as **(c)** positive and **(d)** negative, organized from largest (bottom) to smallest (top) for the MitoD group. Group-specific associations were computed using linear regression models adjusting for age and group-specific percent fat. Circles represent the size of the adjusted bivariate association between FGF21 and the self-report measures as Cohen’s *r* value with 95% CI error bars. Diamonds indicate overall Cohen’s *r* for each group across each category with standard error of mean (SEM). Scatter plots show the three strongest relationships in each category. *Note:* The scale of the Y-axis for FGF21 in the Couples Satisfaction plot differs from the others because Couple’s Satisfaction scores only exist for participants in romantic relationships; MitoD participants with the highest FGF21 levels were not in romantic relationships and did not complete this scale. Two-tailed significance is indicated as *p<*0.05 (*), *p<*0.01 (**), *p<*0.001 (***), *p*<0.0001 (****).

We also analyzed the associations between FGF21 and catecholamines downstream of the sympathetic nervous system (SNS) (i.e., norepinephrine, epinephrine, dopamine, and serotonin; **Figure 2b**). In controls, catecholamines were generally weakly or non-significantly correlated with changing FGF21 levels. By contrast, MitoD individuals with high FGF21 levels tended to have low circulating serotonin and dopamine levels at most timepoints. Although the physiological functions of these neurotransmitters in circulation remains unclear, such differential association patterns reinforce the notion that the FGF21 response to mental stress is moderated by mitochondrial biology.

To interrogate the generalizability of these group-level findings, we leveraged the naturally occurring inter-individual variation in FGF21 responses across our 64 control participants. Stratifying controls based on their FGF21 stress-reactivity (net FGF21 dynamics over the stress time course), we found that 30% of the controls exhibited mild stress-induced increases in FGF21, similar to MitoD participants. Interestingly, these individuals with net positive FGF21 reactivity exhibited different hormonal and metabolic profiles than those with net negative reactivity (the average control group trajectory). Among controls, physiological profiles associated with positive FGF21 stress reactivity included higher cortisol and cortisone reactivity, lower baseline testosterone levels, and higher body fat and insulin levels (**Figure S8**). Cortisol and cortisone are catabolic hormones (energy mobilization by breakdown), whereas testosterone is anabolic (biosynthesis and repair). Thus, this stratified analysis, together with the MitoD vs Control group differences, suggests that mental stress interacts with anabolic-catabolic axes linked to metabolic health to regulate FGF21.

To explore relationships between FGF21 and trait-like psychosocial factors traditionally linked to disease risk or wellbeing, we then examined correlations between FGF21 levels (AM-fasting and PM-fed, separately) and an extensive set of positive and negative psychosocial factors in the MiSBIE cohort (see *Methods*). These measures included *positive factors* such as environmental mastery (i.e., a sense of agency in one’s life), couples’ satisfaction, social support and others; and *negative factors* including loneliness, feeling ineffective at work, neglect during childhood, and others.

In line with the differential regulation of blood FGF21 in response to the acute laboratory psychosocial stressor, the presence of chronic life stressors or the absence of protective factors was consistently more strongly related to FGF21 in MitoD patients than in controls (**Figure 2c-d**). These patterns were most apparent with FGF21 sampled in the fed state (**Figure S9**). Notably, MitoD individuals who experienced more environmental mastery (*r*_s_=-0.44, *p<*0.0001) or more emotional satisfaction in their romantic relationship had lower FGF21 levels (*r*_s_=-0.43, *p*<0.01, **Figure 2c**). By contrast, greater feelings of loneliness were related to higher FGF21 (*r*_s_*=*0.404, *p*<0.0002), but again only in MitoD (**Figure 2d**). These findings suggests that MitoD patients may exhibit greater metabolic vulnerability to psychosocial stressors—again pointing to converging psychobiology linking OxPhos defects and psychosocial factors in relation to FGF21 signaling.

Finally, extending the small MiSBIE cohort, we validated the associations between FGF21 biology and psychosocial factors in 51,818 UK Biobank participants of various health statuses [18] (https://proteome-phenome-atlas.com). Consistent with our couple’s satisfaction result, UKB participants who experienced a marital separation or divorce had elevated levels of plasma FGF21 (*p*=4.3×10^-7^, *β*=0.066, 95% C.I. [0.041 to 0.091], *n*=17,281), after correcting for demographic variables including age, sex and BMI. Financial difficulties were also associated with elevated FGF21 (*p*=6.5×10^-11^, *β*=0.074, 95% C.I. [0.051 to 0.096], *n*=23,320). On the other hand, engaging in non-exercise- and non-alcohol-related leisure and social activities involving connection with others was associated with lower FGF21 levels (*p*=2.7×10^-13^, *β*=-0.065, 95% C.I. [-0.082 to -0.048], *n*=35,860), substantiating a “social” dimension to FGF21 regulation.

Together, these data reveal three main points: 1) FGF21 is acutely regulated by mental stress within minutes in a direction that depends on mitochondrial health, 2) the dynamic association patterns between FGF21 with anabolic/catabolic endocrine axes, and 3) associations between FGF21 and psychosocial factors consistently associated with disease- and age-related outcomes in other medical conditions [19]. Further research is needed to understand how mitochondrial OxPhos capacity and mental stressors converge to regulate FGF21 and support the energetic demands of psychobiological stress responses.

## Data Availability

All data produced in the present study are available upon reasonable request to the authors

https://github.com/mitopsychobio/FGF21_MitoD_Kurade.git

## Acknowledgements

The MiSBIE study was supported by NIH grants R21MH113011, R01MH122706, RF1AG076821, and R01MH137190, the Seed Grant Program for MR Studies of the Zuckerman Mind Brain Behavior Institute at Columbia University, the Robert N. Butler Columbia Aging Center Fellowship Program at the Mailman School of Public Health, the National Center for Advancing Translational Sciences and NIH through grant numbers UL1TR001873, P30CA013696, the Columbia Irving Institute Scholars program, the Wharton Fund, and Baszucki Group to M.P. The authors are grateful to Grace Liu for assistance with the MiSBIE study database.

## Data availability statement

Source data is available on request. MiSBIE study information, detailed protocols, and procedures are available at www.picardlab.org/MiSBIE.

## Author contributions

M.P., R.P.J., C.T. and M.H. designed the MiSBIE study. M.K. performed FGF21 assays and analyzed data. C.K. recruited participants, performed study visits, collected study data, and performed data quality control. N.B.A. analyzed steroid and catecholamine hormones data. C.K. and C.T. analyzed psychosocial questionnaires. A.B. provided statistical guidance and performed the regression models. M.K., Q.C., and M.P. drafted the manuscript. All authors reviewed the final version of the manuscript.

