## Supplemental Figures for "Mitochondrial and Stress-Related Psychobiological Regulation of FGF21 in Humans"

#### Supplemental Figure 1

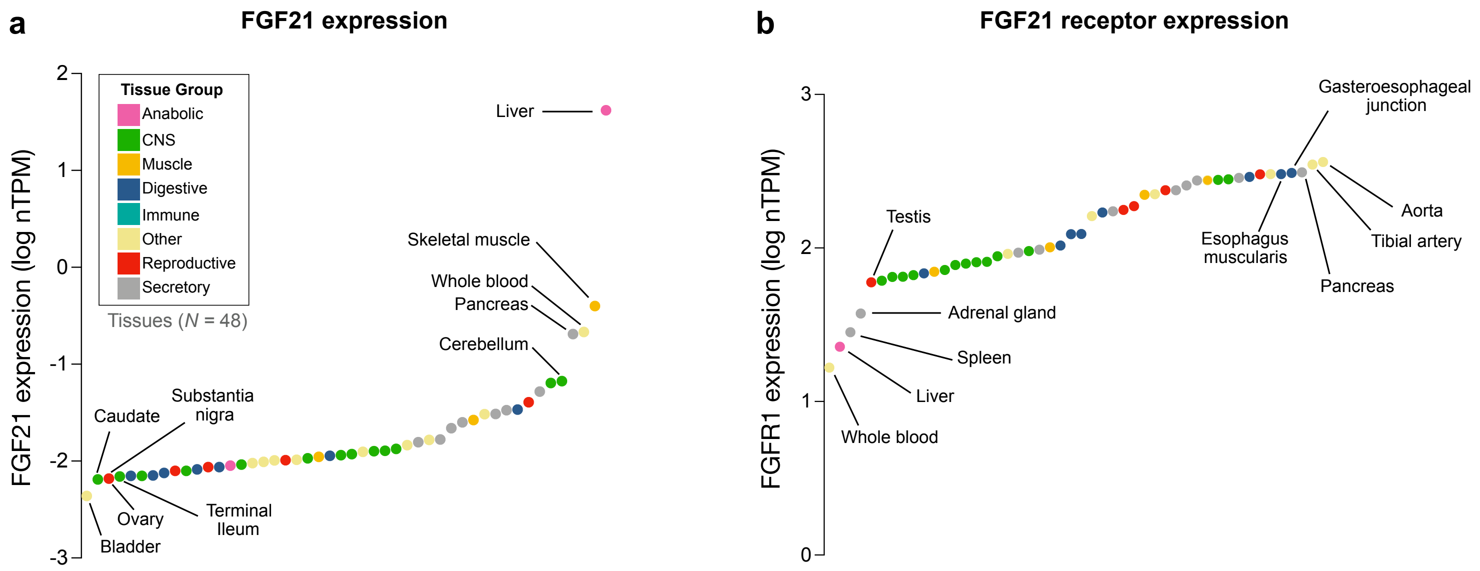


**Supplemental Figure 1. *FGF21* and *FGFR1* expression across the human body.**
Mean transcript levels (normalized Transcript Per Million, nTPM) for (**a**) *FGF21* and its (**b**) receptor *FGFR1* across 48 human tissues, from the GTEx v8 RNAseq dataset. Datapoints represent the mean from 21 to 803 individuals per tissue. Tissues are grouped and color-coded by their systemic functions and location. The five tissues with the highest and lowest expression are annotated. See **Supplemental Table 1** for corresponding values. Wilcoxon signed rank tests showed that liver *FGF21* expression is 104.2-fold higher ($p < 0.0001$) than the second highest tissue, skeletal muscle, and 123.7-fold higher than the average of all human tissues. *Note:* The relatively uniform expression of *FGFR1* across vascular, digestive, and glandular tissues, and two brain regions is consistent with circulating FGF21 having a broad signaling in human physiology.

### Supplemental Figure 2

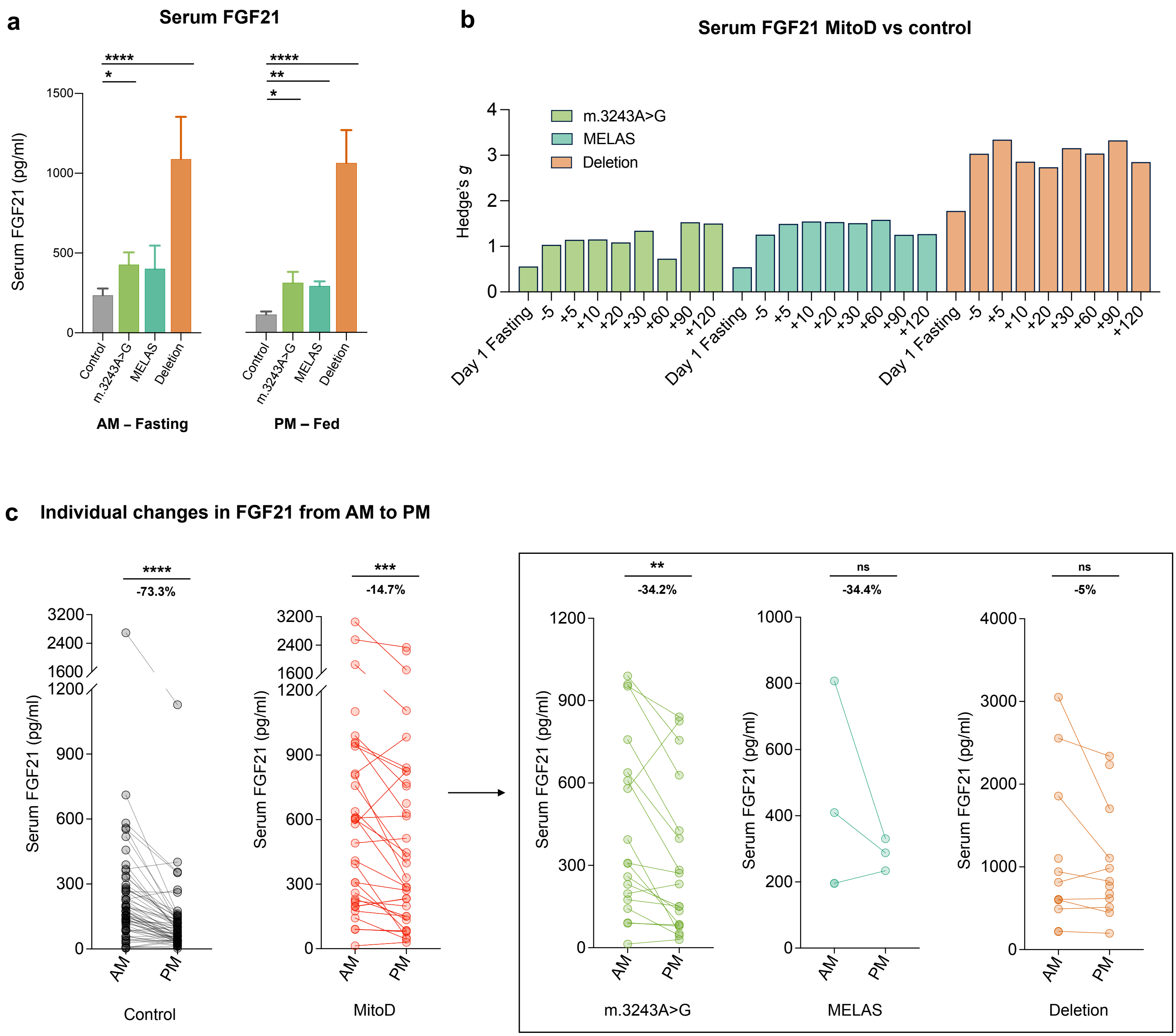


**Supplemental Figure 2. Morning to Afternoon Dynamics of FGF21 Across Groups.**
**(a)** Mann-Whitney's U-test comparing fasting and fed mean FGF21 levels (pg/ml) between controls (*n* = 65) and MitoD subgroups (3243A>G, *n* = 18; MELAS, *n* = 3-4; Deletion, *n* = 12). In the AM–Fasting state, the 3243A>G group was 0.8-fold higher (*M* = 427.6 pg/ml, *p* = 0.0005) than controls (*M* = 235 pg/ml), whereas the Deletion group was 3.6-fold higher (*M* = 1088 pg/ml, *p* < 0.0001). In the PM–Fed state, all MitoD groups were significantly higher than controls (*M* =109 pg/ml): 3243A>G was 1.8-fold higher (*M* = 304 pg/ml, *p* = 0.0005), MELAS was 1.6-fold higher (*M* = 284 pg/ml, *p* = 0.005), and Deletion was 8.5-fold higher (*M* = 1035 pg/ml, *p* < 0.0001). **(b)** Hedge's *g* comparing MitoD subgroups to controls at each time point. **(c)** Fasting (AM) to fed (PM) change in individual participants' FGF21 (pg/ml) levels with the group difference expressed as percent difference between time points. Paired Wilcoxon tests showed significant decreases from fasting-AM to fed-PM levels in controls (*n* = 61, *p* < 0.0001, Median: AM = 173.8 pg/ml, PM = 79.08 pg/ml) and the MitoD group combined (*n* = 30, *p* = 0.0005, Median: AM = 535.4 pg/ml, PM = 309.1 pg/ml). When split by subgroup, there was a significant decrease in the m.3243A>G point mutation group only (*n* = 18, *p* = 0.004, Median: AM = 307 pg/ml, PM = 191.6 pg/ml).
*p* < 0.05 (*), *p* < 0.01 (**), *p* < 0.001 (***), *p* < 0.0001 (****).

#### Supplemental Figure 3

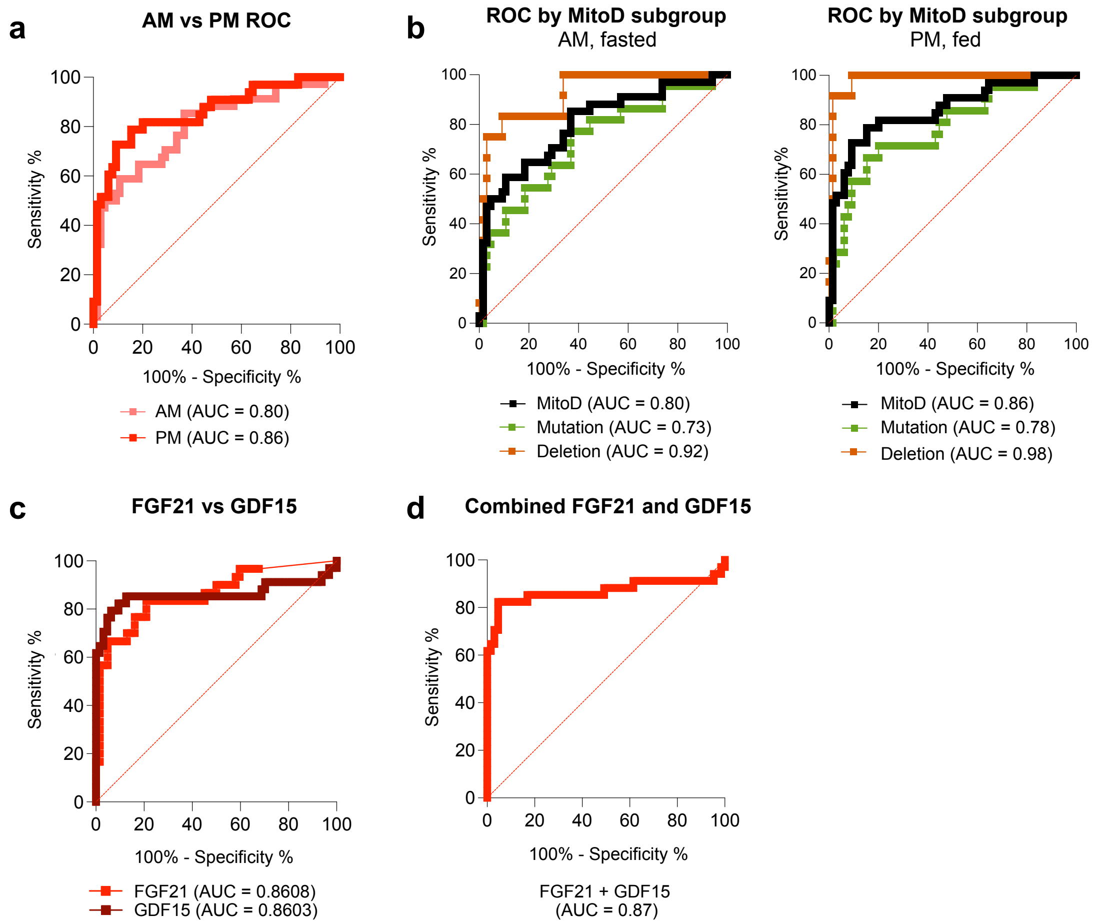


**Supplemental Figure 3. ROC Analyses examining FGF21 as a Diagnostic Biomarker for Mitochondrial Disease.** Receiver operating characteristic (ROC) curve analyses for the prediction of mitochondrial disease based on the serum levels of FGF21 measured by enzyme-linked immunosorbent assay. Plots depict percentage sensitivity plotted against percentage specificity to assess diagnostic performance of serum FGF21 in distinguishing MitoD from healthy participants. **(a)** Two curves comparing the diagnostic performance of fasting (AM) versus fed (PM) measures of serum FGF21, showing that fed (AM) FGF21 shows enhanced performance with AUC = 0.86. **(b)** Two plots comparing the diagnostic performance of fed and fasting FGF21 for Deletion and Mutation (m.3243A>G + MELAS) groups separately, alongside a groups-combined curve. Curves demonstrate highest discriminative ability for the Deletion group in the fed state (AUC = 0.98). **(c)** Comparison of FGF21 and GDF15 diagnostic performance in the fed state and **(d)** the performance of FGF21 and GDF15 combined. The combined curve was generated using individual FGF21 and GDF15 measures and calculating predicted probabilities by logistic regression for each participant measurement at PM-fed time point.
*p* > 0.05 (ns), *p* < 0.05 (*), *p* < 0.01 (**), *p* < 0.001 (***), *p* < 0.0001 (****).

### Supplemental Figure 4

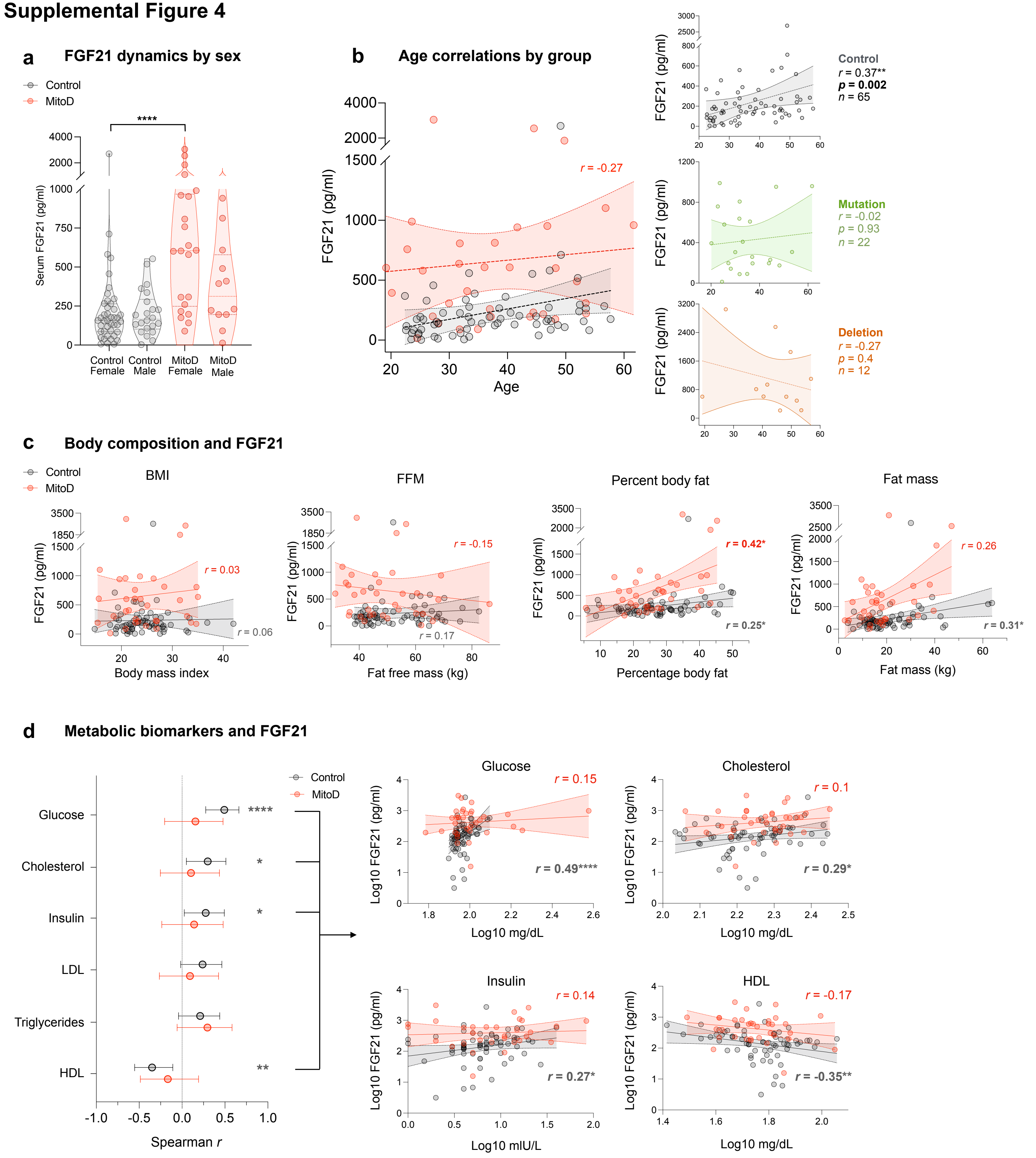


**Supplemental Figure 4. Demographic and metabolic variations in serum FGF21 levels by group: Analysis by sex, age, body composition and metabolic markers. (a)** A Dunn's multiple comaparison test revealed no difference in FGF21 levels between females (*n* = 64) and males (*n* = 34). **(b)** Spearman correlations between age and fasting FGF21 (pg/ml) by group. **(c)** Spearman correlations between fasting FGF21 levels (pg/ml) and different body composition factors in controls and MitoD participants. Percent body fat was significantly correlated with FGF21 in both groups (controls, *n* = 63, *p* = 0.04, and MitoD, *n* = 34, *p* = 0.01), whereas fat mass was only significantly correlated in controls (*n* = 63, *p* = 0.01). **(d)** Forrest plot depicting Spearman correlations between fasting FGF21 and metabolic biomarkers in controls and MitoD participants; all measurements are Log10 transformed and error bars represent 95% confidence intervals. In controls (*n* = 65), FGF21 was positively correlated with blood glucose ($p < 0.0001$), total cholesterol ($p = 0.01$), and insulin ($p = 0.03$), and negatively correlated with HDL ($p = 0.004$). The MitoD group showed no significant correlation with any metabolic biomarkers.

$p < 0.05$ (*), $p < 0.01$ (**), $p < 0.001$ (***), $p < 0.0001$ (****).

#### Supplemental Figure 5

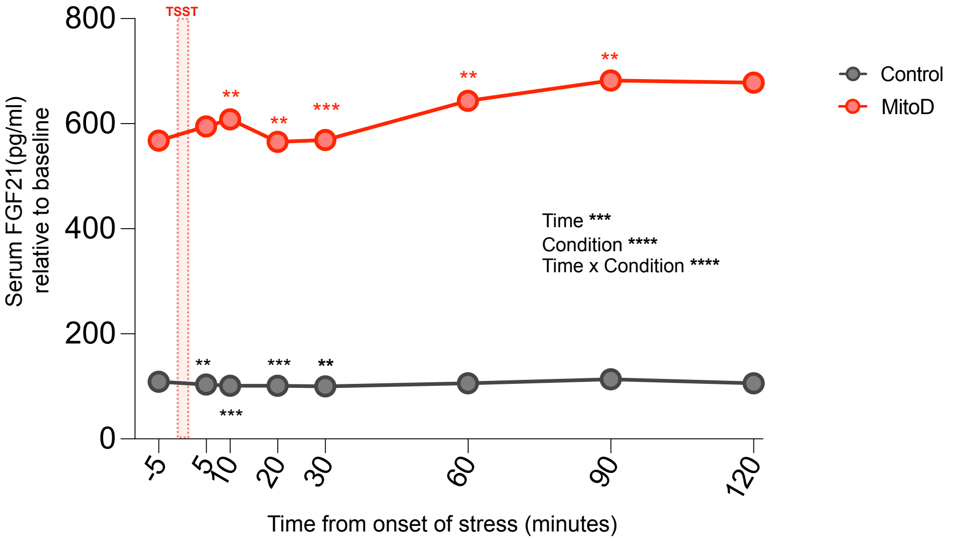


**Supplementary Figure 5. Mixed-effects model comparing MitoD to controls over time.** A mixed effects model with Restricted Maximum Likelihood (REML) estimation showed significant effects of Group, $F(1,100) = 44.34$, $p < 0.0001$, Time, $F(1.2,109.9) = 13.05$, $p = 0.0002$, and a significant Group X Time interaction, $F(7, 640) = 12$, $p < 0.0001$, indicating that the trajectory of serum FGF21 over time varied significantly in the MitoD group compared to controls. Using Dunnett's tests to compare each subsequent time point to baseline (-5min), we found that controls showed significant differences at 5mins ($p = 0.0021$), 10mins ($p = 0.0003$), 20mins ($p = 0.0007$), and 30mins ($p = 0.0014$), whereas the MitoD group showed significant differences at 10mins ($p = 0.0091$), 20mins ($p = 0.006$), 30mins ($p = 0.0004$), 60mins ($p = 0.0013$), and 90mins ($p = 0.0052$).
$p < 0.05$ (*), $p < 0.01$ (**), $p < 0.001$ (***), $p < 0.0001$ (****).

### Supplemental Figure 6

**a** Baseline corrected serum FGF21 (pg/ml) in controls

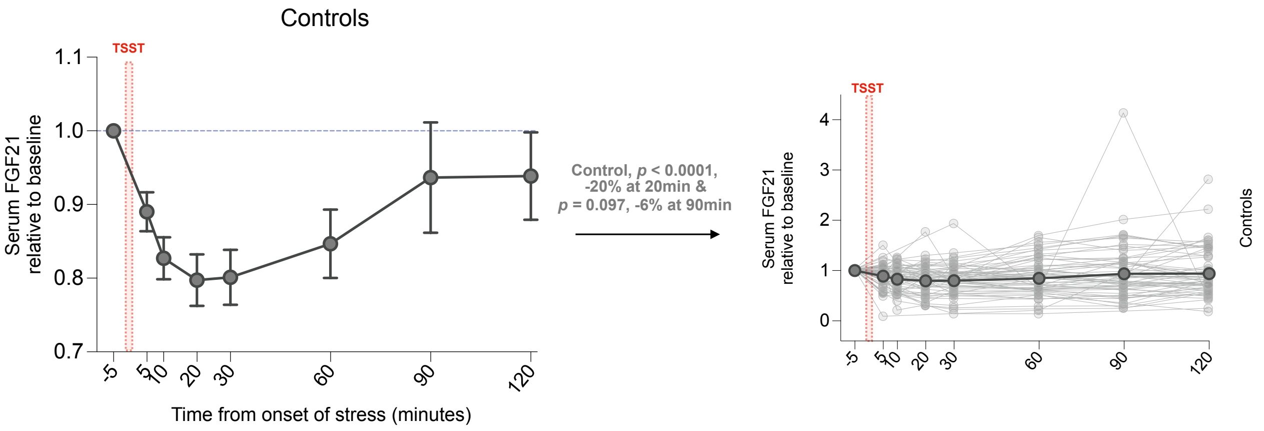


**b** Baseline corrected serum FGF21 (pg/ml) in all MitoD groups

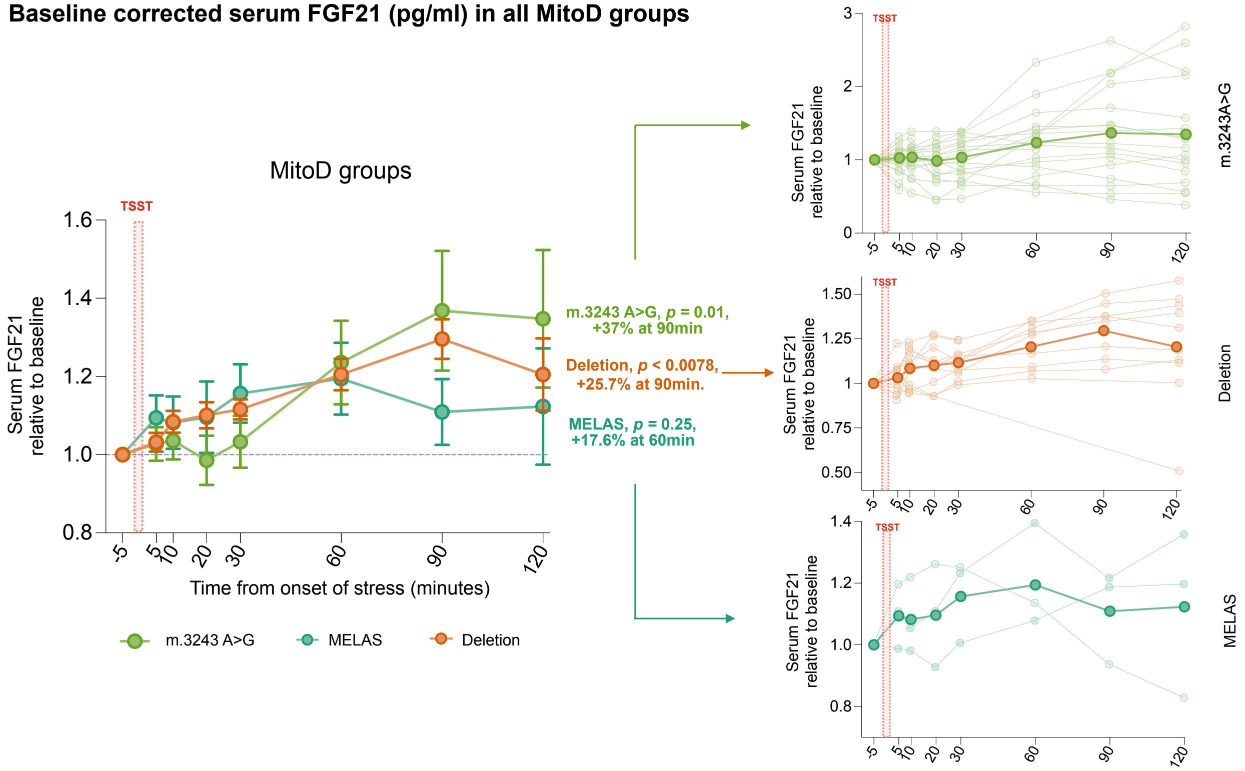


**Supplemental Figure 6. Individual and group FGF21 trajectories in response to the speech task.**
Individual and group trajectories for each group are show relative to baseline values (Value/Baseline). Relative change between time points was calculated as the value at a given time point minus the baseline value (-5min). For each group, paired Wilcoxon tests were used to compare the FGF21 peak to the group baseline **(a)** Average percent change in FGF21 (pg/ml) in controls as a group, as well as individual trajectories for each participant. **(b)** Average percent change for each MitoD subgroup relative to their baseline, as well as individual trajectories for each participant.

#### Supplemental Figure 7

a

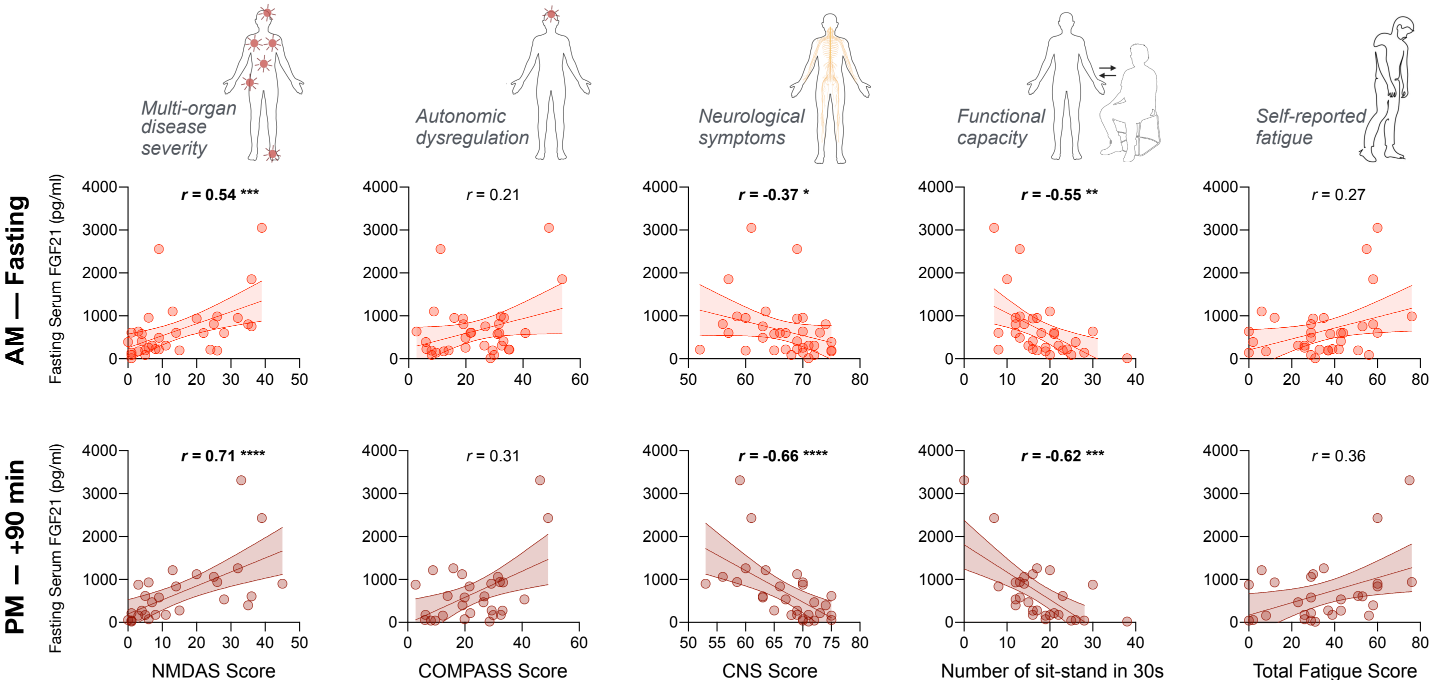


b

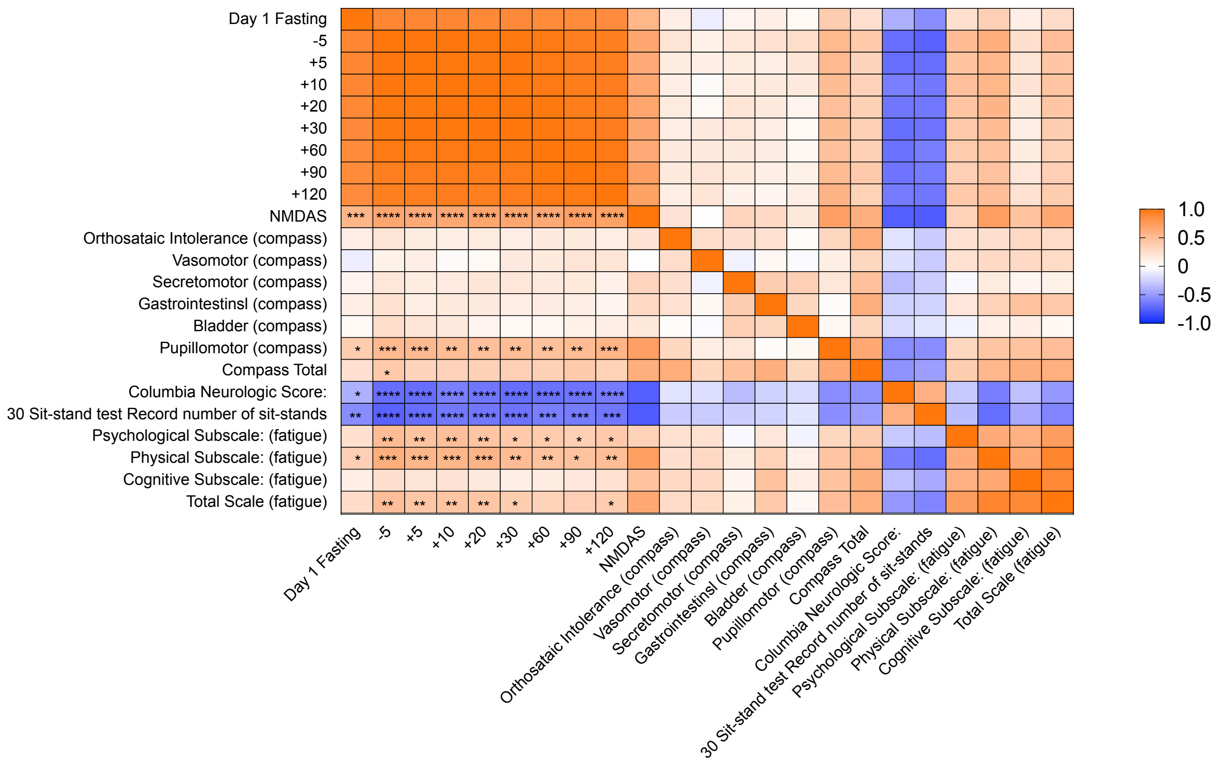


**Supplemental Figure 7. Psychosocial stress enhances correlations between FGF21 and disease severity indices in patients with MitoD.** **(a)** Scatter plots illustrating Spearman correlations between FGF21 and composite scores of various disease severity indices of at fasting (AM) and post-stress (+90min). **(b)** Spearman correlation matrix showing correlations between serum FGF21 levels across time points and subscales of disease severity indices. NIMDAS = The Newcastle Mitochondrial Disease Adult Scale.
*p* ≤ 0.05 (*), *p* ≤ 0.01 (**), *p* ≤ 0.001 (***), *p* ≤ 0.0001 (****).

### Supplemental Figure 8

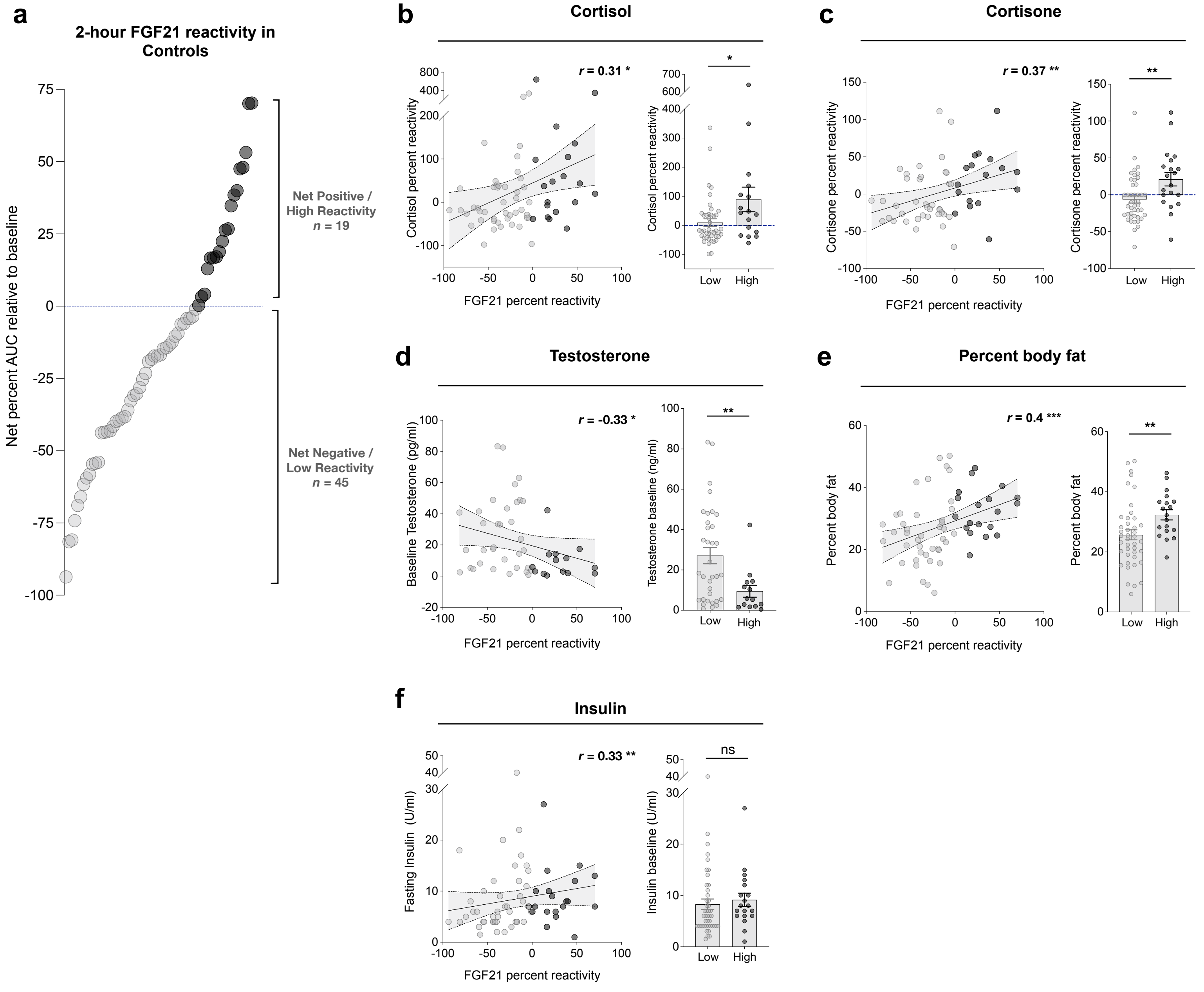


**Supplemental Figure 8. Differences in controls based on FGF21 reactivity.**
**(a)** Controls classified as having a net positive or net negative FGF21 reactors based on percent Area Under the Curve (AUC). AUC calculations were derived from baseline corrected values expressed as percent change relative to baseline FGF21, with both positive and negative peaks included in this calculation, resulting in a net percent AUC for each participant. Scatter plots show significant Spearman correlations between FGF21 reactivity and **(b)** cortisol reactivity ($p$ = 0.012, $n$ = 64), **(c)** cortisone reactivity ($p$ = 0.0049, $n$ = 55), **(d)** baseline testosterone levels ($p$ = 0.019, $n$ = 48), **(e)** percent body fat ($p$ = 0.0005, $n$ = 61), and **(f)** fasting insulin levels ($p$ = 0.0069, $n$ = 64). Reactivity for cortisol and cortisone was calculated as Percent Reactivity = ((Max_Reactivity_Value - Baseline_Value) / Baseline_Value) * 100; Baseline testosterone = measurements (pg/ml) at -5min of the TSST time course; Fasting insulin = measurements (U/ml) at the fasting (AM) time point. Bar graphs for unpaired Mann-Whitney's tests show that controls with positive reactivity have **(b)** 80% higher mean cortisol reactivity ($p$ = 0.033, $n$ = 43 vs 17), **(c)** 30% higher mean cortisone reactivity ($p$ = 0.0023, $n$ = 37 vs 18), and **(e)** 6.6% higher mean percent body fat ($p$ = 0.0056, $n$ = 42 vs 19). **(d)** Inversely, controls with higher FGF21 reactivity had 17.61 pg/ml lower mean testosterone at baseline ($p$ = 0.0066, $n$ = 34 vs 14). **(f)** There was no statistically significant difference between high and low FGF21 reactors on baseline insulin ($p$ = 0.16, $n$ = 45 vs 19).
$p$ > 0.05 (ns), $p$ < 0.05 (*), $p$ < 0.01 (**), $p$ < 0.001 (***), $p$ < 0.0001 (****).

#### Supplementary Figure 9

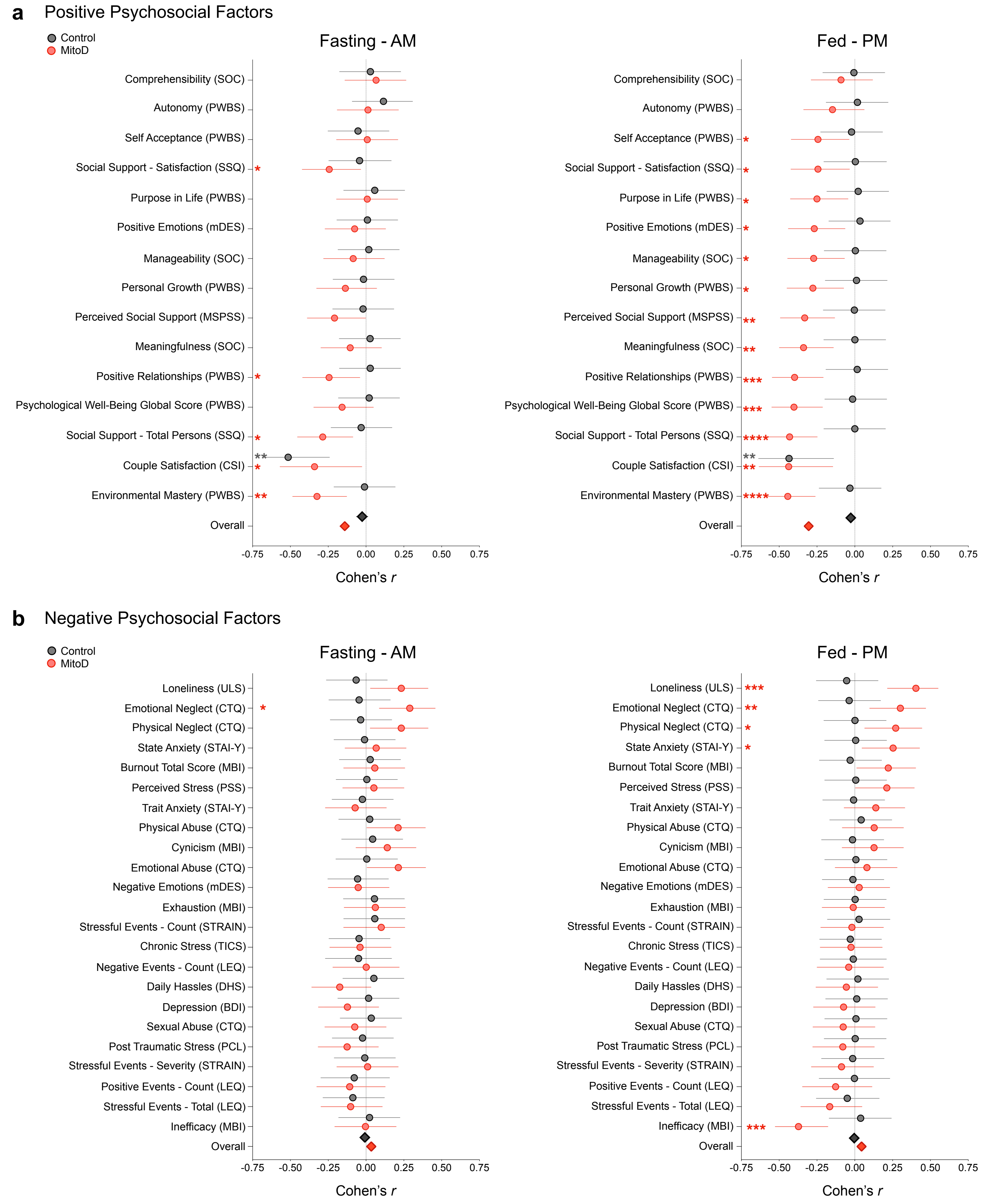


**Supplemental Figure 9. Forrest plots of fasting and fed FGF21 levels with psychosocial self-report measures.** Forrest plots illustrating correlations between Fasting and Fed FGF21 levels, adjusted for age and percent fat, with psychosocial self-report measures categorized as **(a)** positive or **(b)** negative. Fasting is presented on the left and Fed on the right, with the comparison showing that the differences between controls and MitoD are more apparent in the Fed state. The group-specific associations were computed in linear regression models adjusting for age and group-specific percent fat. Circles represent the size of the adjusted bivariate association between FGF21 and the self-report measures as Cohen's r value with 95% CI error bars. They are organized from smallest (bottom) to largest (top) correlation for the MitoD group in the Fed state. Diamonds represent the overall Cohen's *r* for each group represented as mean with Standard Error of Mean (SEM). *p* < 0.05 (*), *p* < 0.01 (**), *p* < 0.001 (***), *p* < 0.0001 (****).
