## Supplemental Materials for "Mitochondrial and Stress-Related Psychobiological Regulation of FGF21 in Humans"

### Methods

#### Participants

Participants were enrolled in the Mitochondrial Stress, Brain Imaging, and Epigenetics (MiSBIE) study in accordance with the guidelines set by the New York State Psychiatric Institute IRB protocol #7424 and the Columbia University Medical Center IRB protocol #AAAU9470. The study was registered in clinicaltrials.gov under #NCT04831424. Participants were recruited locally from our clinic at the Columbia University Irving Medical Center and across the United States and Canada. All participants provided written informed consent to participate in the study and authorizing the potential dissemination of results. Recruitment took place between June 2018 and May 2023. A total of 110 participants aged 18 to 60 years were enrolled, including 70 healthy controls (*n* = 48 female, 22 male) and 40 individuals with genetically defined mitochondrial diseases (MitoD, *n* = 28 female, 12 male). Eligibility criteria can be found in [1].

#### Procedure

The fasting assessments and psychosocial stress paradigm described here were part of an extended 2-day MiSBIE study protocol described in detail in [1]. Day 1 included a fasting blood draw in the morning followed by a clinical assessment, completion of self-report questionnaires, and an extensive psychophysiological protocol that included a modified Trier Social Stress Test (TSST)—a well-established method for inducing and measuring psychosocial stress responses [2].

The modified TSST speech task administered in this study consisted of 120 seconds of speech preparation followed by 180 seconds of speech delivery under evaluation. To induce stress, the study coordinator entered the room carrying a clipboard and a video camera mounted on a tripod. The coordinator silently set up the video camera to give the impression that it was recording, and placed a full-length mirror in front of the participants so they could see themselves throughout the task. Participants were instructed to prepare a speech in response to an uncomfortable scenario: being accused by a security guard of stealing merchandise from a store. After the instructions and scenario were presented, participants completed a subset of items from Primary Appraisal Secondary Appraisal (PASA) threat questionnaire. They were given two minutes to prepare their speech and were then instructed to deliver it to an evaluator for three minutes. The “evaluator”, also wearing a white coat and carrying a clipboard, entered the room silently and took notes as the participants delivered their speeches. After the threeminute period, the evaluator left and the camera and mirror were removed simultaneously. Blood and saliva samples, body temperature, and affect ratings were collected 5 minutes before the initiation of the speech task, and again 5, 10, 20, 30, 60, 90, and 120 minutes post-TSST.

### Sample Collection and Processing

#### *Blood*

Blood was collected by an intravenous catheter in the antecubital vein at nine different timepoints. The fasting sample was collected in the morning of Day 1 (2 x 8.0ml serum clot activator tubes [BD-367820]) and eight afternoon samples were collected (8 x 6.0ml serum clot activator tubes [BD-367815]) during the stress reactivity protocol of Day 1. Samples were inverted 10-12 times and allowed to rest for at least 30 minutes before serum isolation. Serum was isolated by centrifugation and aliquoted to be stored at -80°C until used. Minor deviations between the handling of the morning and afternoon samples can be found in the detailed blood processing protocol reported in [1].

#### *Saliva*

Saliva was collected with a cotton salivette (Sarstedt, Cat# 51.1534.500, Nümbrecht, Germany) at 9 timepoints, including the Day 1 morning sample and the 8 afternoon samples collected during the stress reactivity protocol. Collected samples were placed on ice and transported to the lab immediately (morning) or were stored on ice until the end of the protocol (afternoon) and then transferred to the laboratory where the salivettes were centrifuged at 1000g for 5 minutes at 4°C. The supernatant was carefully aliquoted into 2ml cryogenic storage tubes and stored in -80 freezer immedeiately. Minor deviations between the handling of the morning and afternoon samples can be found in the detailed blood processing protocol reported in [1].

### Measures

#### *FGF21*

FGF21 levels were measured in serum using a high-sensitivity ELISA kit (R&D Systems, DF2100) according to the manufacturer's instructions. Similar lot numbers were employed throughout all participant samples, and the average coefficient of variation (CV) between lot numbers was determined using reference samples for quality control. Serum samples were diluted with the calibrator diluent provided in the kit at a 1:4 ratio. Absorbance was measured at two wavelengths, 450nm and 540nm. Optical imperfections in the plate were corrected by subtracting readings at 540nm from the readings at 450nm. Samples were analyzed on duplicate plates, with concentrations calculated as the average of the duplicates. A standard curve (5 samples per plate) and serum reference samples (3 samples per plate, the same sample per batch) were run with each assay, and the inter-assay CV was monitored. All standard curves and references were overlaid to detect failed runs. Samples at all concentration readings were included in the final analysis, as the control cohort had low detection rates. For samples with concentrations below the standard curve, adjustments were made by calibrating the values to half of the minimum standard curve concentration. Despite this calibration of low concentrations, the significance of the various analyses performed was not affected. After confirming that adjusting the values for low detection had no impact on the results, all original concentration values were used in the analysis.

Across the entire study cohort, we had total 131 missing samples due to failed blood draws (e.g., veins too small) or technical issues in processing samples. There were 11 missing samples for the fasting timepoint and a total of 120 across the stress time course. For controls, there were 5 missing samples at both fasting and the baseline stress (-5 min) timepoints. For the MitoD group, there were 6 missing samples at fasting and 7 missing samples for baseline.

***Catecholamine Hormones***

Norepinephrine, Epinephrine, Dopamine, and Serotonin were measured in serum using Liquid Chromatography-Tandem Mass Spectrometry (UPLC-MSMS) by the Biomarker Core Laboratory at Columbia University's Irving Institute for Clinical and Translational Research. The metabolites were extracted from samples spiked with deuterated internal standards via protein precipitation, followed by derivatization with dansyl chloride. The supernatant was then subjected to liquid-liquid extraction using ethyl acetate, and the extracted metabolites were resuspended in acetonitrile for LC-MS analysis. Chromatographic separation was achieved on a Waters ACQUITY UPLC HHS C18 column (2.1 × 100 mm, 1.8 µm) maintained at 40°C, with gradient elution using water and acetonitrile containing 0.1% formic acid as mobile phases at a flow rate of 300 µL/min. LC-MS/MS analysis was performed in positive ESI mode with multiple reaction monitoring (MRM) (transitions: norepinephrine: 869.2>170.3; epinephrine: 883.3>170.2; dopamine: 853.30>170.37; serotonin: 643.29>146.05) on a Waters Xevo TQS MS integrated with the ACQUITY UPLC system (Waters, Milford, MA, USA).

***Steroid Hormones***

Cortisol, cortisone, DHEA, and testosterone levels were measured in saliva samples by a high-performance liquid chromatography–tandem mass spectrometry (LC–MS/MS) using API 5000 QTrap mass spectrometer (AB Sciex#6669), following previously established methods [3, 4]. Previously aliquoted saliva samples were thawed, and 100 µL of saliva was added to a tube containing 50 µL of internal standard and 100 µL of a methanol/water solution with zinc sulfate. The mixture was vortexed, centrifuged, and the supernatant ant was injected into the LC-MS/MS system for analysis. Cortisol, cortisone, and testosterone were detected within a range of 0.001-10 ng/mL, and DHEA within a range of 0.01-20 ng/mL, with intra- and inter-assay coefficients of variation between 4.3% and 10.8%.

Across the entire study cohort, out of 880 samples for catecholamine and steroid measurements, the dataset included a total of 72 missing samples for cortisol, 73 for cortisone, 267 for testosterone, 547 for DHEA, 445 for epinephrine, 116 for norepinephrine, 116 for dopamine, 116 for serotonin. Data is missing either because saliva/blood could not be collected or because the analyte concentration was lower than the detectable limit.

***Metabolic Markers***

Glucose, insulin, total cholesterol, high-density lipoprotein, low-density lipoprotein, and triglycerides levels were measured from fasting samples by a CLIA-certified diagnostic laboratory at Columbia Presbyterian Hospital according to their standard protocols.

***Body Composition Markers***

Participants height and weight were measured on the morning of Day 1 and Body Mass Index was calculated as BMI = weight [kg]/height $[m]^2$. Percent fat mass and fat-free mass (FFM) were estimated using a 4-point bioelectrical impedance device (InnerScan PRO, Multi-Frequency, Segmental, Wireless, Body Composition Monitor; Tanita, SKU:MT05, Arlington Heights, IL).

***Disease Severity Markers***

Disease severity and symptom profiles were evaluated using five different methods: 1) the Newcastle Mitochondrial Disease Adult Scale (NMDAS)—a validated tool to assess symptoms and functional limitations across various life domains and the 2) the Columbia Neurological Score (CNS) used to measure neurological symptoms focused on the central nervous system (both of which are administered by a qualified clinician); 3) the Composite Autonomic Symptom Score (COMPASS-31), which evaluates autonomic dysregulation across six domains; 4) the 30-second sit-to-stand test to measure functional capacity; and 5) the Modified Fatigue Impact Scale (MFIS) to measure perceived fatigue and its effects on physical, cognitive, and psychosocial functioning [5].

***Psychosocial Self-report measures***

Aspects of participants' psychosocial lives were measured using a battery of standardized self-report measures, which included the following domains:

**Perceived Stress and Stress Exposure**

The dimensions of perceived stress and stress exposure assessed included perceived stress measured with the Perceived Stress Scale (PSS) [6], chronic stress measured with the Tried Inventory for the Assessment of Chronic Stress (TICS) [7], daily hassles measured with the Daily Hassles Scale (DHS) [8], childhood trauma measured with the Childhood Trauma Questionnaire (CTQ) [9], life events measured with the Life Events Questionnaire (LEQ) [10], and lifetime stressor exposure measured with the Stress and Adversity Inventory for Adults (STRAIN) [11].

**Mental Health and Wellbeing**

The dimensions of health and well-being assessed included anxiety measured with the State and Trait Anxiety Inventory (STAI-Y) [12], depressive symptoms measured with the Beck Depression Inventory (BDI) [13], burnout measured with the Maslach Burnout Inventory (MBI) [14], post-traumatic stress disorder (PTSD) symptoms measured with the PTSD Checklist for the DSM-5 (PCL-C) [15], personal well-being measured with Ryff's Psychological Well-Being Scale (PWBS) [16], aspects of an individual's sense of coherence measured with Antonovky's Sense of Coherence Scale (SOC) [17], and daily emotions measured with the Modified Differential Emotions Scale (mDES) [18].

**Social Life**

The dimensions of social life assessed included perceived social status measured with the MacArthur Ladder [19], social support measured with the Social Support Questionnaire (SSQ) [20] and the Multidimensional Scale of Perceived Social Support (MSPSS) [21], couple satisfaction measured with the Couple Satisfaction Index (CSI) [22], and loneliness measured with the UCLA Loneliness Scale Version III (ULS) [23].

### Statistical analysis

Spearman's rank correlations were computed on raw values with exceptions for catecholamines/steroid hormones and metabolic biomarkers, which were log transformed. Group differences among controls and MitoD cohorts, stratified by sex and individual group differences between the control, mutation, and deletion groups were evaluated using the non-parametric Mann-Whitney *U*-tests and Kruskal-Wallis tests, followed by Dunn's multiple-comparison test for post-hoc analysis when appropriate. Differences between two timepoints in the same individuals (e.g., morning to afternoon, Day 1 fasting to fed) were analyzed using the non-parametric Wilcoxon signed-rank test for paired data.

Stress reactivity was defined as either relative or absolute change from baseline at each time point. To determine whether acute psychosocial stress affected serum FGF21 levels over time, and if these results differed between groups (MitoD vs. controls), we used a linear mixed effects model to test the effects of Group, Time, and the Group x Time interaction. Because the assumption of sphericity was violated, the Geisser and Greenhouse correction was applied. Post-hoc Dunnett's tests were performed for multiple within-group comparisons to test FGF21 stress reactivity within each group. Hedge's *g* was used as a measure of effect size. Correlations, Wilcoxon tests, the mixed effects model, and Dunnett's tests were performed using GraphPad 10.0 (GraphPad Software, San Diego, California, USA).

Associations between FGF21 levels and psychosocial self-report measures were computed using linear regression models in R (version 2023.12.1+402). An interaction term of the respective psychosocial self-report measures with group (MitoD vs. controls) served to compute the disease group-specific associations between FGF21 and the psychosocial self-report measures. All models were adjusted for the covariates age and percent fat due to their relevant influence on FGF21 (**Supplementary Figures 3 and 4**). The adjusted bivariate association between FGF21 and the self-report measures was computed and tested for significance per disease group using simple slope analyses (R package *emmeans*), reporting Cohen's *r* as a standardized effect size measure.

### FGF21 and psychosocial associations from the UK Biobank plasma proteome atlas

Finally, we compared our MiSBIE results with FGF21 findings from the atlas of plasma proteins (https://proteome-phenome-atlas.com/) generated from the UK Biobank, which examined 2,920 plasma proteins in relation to 986 health-related traits in 53,026 individuals [24]. Regression models were conducted by the 'lm', 'glm' and the 'polr' function from the R package

‘MASS’ (v4.2.0). Associations between plasma proteins and binary health-related traits (e.g., exposures) were assessed via logistic regression, and associations with ordered categorical traits were assessed with proportional odds logistic regression models. All regressions were performed with the adjustment of participants’ baseline information of age, sex, ethnicity, Townsend deprivation index, Body-Mass Index (BMI), smoking status, fasting time, season of blood collection (summer/autumn: June to November versus winter/spring: December to May) and blood age (date of blood collection to date of protein examination). Significant associations were defined with a threshold of $p_{adj} < 1.71 \times 10^{-8}$ ($p < 0.05$ / [2,920∗approximately 1,000 traits]) to correct for multiple tests. Adjusted $p$ values, $\beta$, 95% confidence intervals, and sample sizes are reported in the text.
